# Structured Clinical Reasoning Prompt Enhances LLM’s Diagnostic Capabilities in Diagnosis Please Quiz Cases

**DOI:** 10.1101/2024.09.01.24312894

**Authors:** Yuki Sonoda, Ryo Kurokawa, Akifumi Hagiwara, Yusuke Asari, Takahiro Fukushima, Jun Kanzawa, Wataru Gonoi, Osamu Abe

## Abstract

**Background:** Large Language Models (LLMs) show promise in medical diagnosis, but their performance varies with prompting. Recent studies suggest that modifying prompts may enhance diagnostic capabilities.

**Objective:** This study aimed to test whether a prompting approach that aligns with general clinical reasoning methodology—specifically, separating processes of summarizing clinical information and making diagnoses based on the summary instead of one-step processing—can enhance LLM’s medical diagnostic capabilities

**Methods:** 322 quiz questions from *Radiology’s* Diagnosis Please cases (1998-2023) were used. We employed Claude 3.5 Sonnet, a state-of-the-art LLM, to compare three approaches: 1) Conventional zero-shot chain-of-thought prompt, as a baseline, 2) two-step approach: LLM organizes patient history and imaging findings, then provides diagnoses, and 3) Summary-only approach: Using only the LLM-generated summary for diagnoses.

**Results:** The two-step approach significantly outperformed both baseline and summary-only methods in diagnosis accuracy, as determined by McNemar tests. Primary diagnosis accuracy was 60.6% for the two-step approach, compared to 56.5% for baseline (p=0.042) and 56.3% for summary-only (p=0.035). For the top three diagnoses, accuracy was 70.5%, 66.5%, and 65.5% respectively (p=0.005 for baseline, p=0.008 for summary-only). No significant differences were observed between baseline and summary-only approaches.

**Conclusion:** Our results indicate that a structured clinical reasoning approach enhances LLM’s diagnostic accuracy. This method shows potential as a valuable tool for deriving diagnoses from free-text clinical information. The approach aligns well with established clinical reasoning processes, suggesting its potential applicability in real-world clinical settings.

## Introduction

The rapid advancement of LLMs has sparked considerable interest in their potential applications across various fields, with medicine being a promising area (1). These models have demonstrated capabilities that extend beyond simple tasks such as explanation and dialogue, showcasing impressive abilities in reasoning and analysis (2). In the medical domain, a large number of studies have already provided evidence of LLMs’ clinical reasoning capabilities (3,4,5). For instance, in the field of diagnostic radiology, Ueda et al. showed OpenAI’s GPT-4 model correctly answered 170 out of 313 cases in “Diagnosis Please,” a monthly diagnostic radiology quiz case series for radiology experts published in the international academic journal *Radiology* (6).

Effective use of LLMs relies on prompt engineering. Studies have suggested that the performance of these models in reasoning tasks can be enhanced by encouraging them to articulate intermediate steps. This approach, referred to as zero-shot chain-of-thought (CoT) prompting, has shown promising results across various domains (7). Savage et al. compared traditional CoT prompting with four clinical reasoning strategies: differential diagnosis, intuitive reasoning, analytical reasoning, and Bayesian inference (4). They reported that GPT-4 showed no significant decrease in performance with these strategies and suggested that this approach could contribute to improving the interpretability of the model’s outputs. Wada et al. demonstrated that by prompting the LLM to output its confidence in its diagnoses, and using these values as thresholds, they were able to reduce the false positive rate of the LLM (8). Fukushima et al. showed that by explicitly indicating within the prompt that the cases being addressed were from a medical journal’s quiz series, the diagnostic accuracy improved, while erroneously informing in the prompt that the setting was primary care resulted in decreased accuracy (9).

Although much remains unknown about LLM functionality and prompt engineering, techniques inspired by human cognitive processes show promise in various applications. Zhou et al. demonstrated that when having LLMs handle various tasks, dividing these problems into simpler and more manageable sub-problems can improve performance (10). Following this line of reasoning, we hypothesized that LLMs would perform better in clinical reasoning tasks when prompted to systematically list and organize patient information (e.g., medical history, family history, lifestyle factors) from free-text clinical descriptions, mirroring the approach of human clinicians (11).

This study aims to evaluate the impact of an explicit intermediate prompt where LLM summarizes clinical information on LLMs’ diagnostic reasoning performance. Additionally, if there were differences in diagnostic performances resulting from this, we investigated whether these differences were due to the summarized information itself or the combination of the original text and the structured summary by comparing the diagnostic accuracies provided by three types of prompting approaches: conventional zero-shot chain-of-thought prompt, two-step summarizing approach for patient information, and summary-only approach.

## Methods

### An overview of this study is presented in Fig. 1

We utilized Claude 3.5 Sonnet, a commercially available LLM developed by Anthropic, to list the primary diagnosis and two differential diagnoses for the 322 consecutive quiz questions (cases 1–322, published between 1998 and 2023) from *Radiology’s* Diagnosis Please (https://dxp.rsna.org/). This specific model was selected due to its superior performance in quiz-based clinical reasoning tasks in the field of diagnostic radiology, as evidenced by recent comparative studies (12,13). The model was accessed through its application programming interface (claude-3-5-sonnet-20240620, accessed on Aug 18, 2024). The model parameters were configured with a temperature setting of 0, as previous studies have suggested that this setting does not negatively impact accuracy while ensuring better reproducibility of the results (14).

**Fig. 1.**
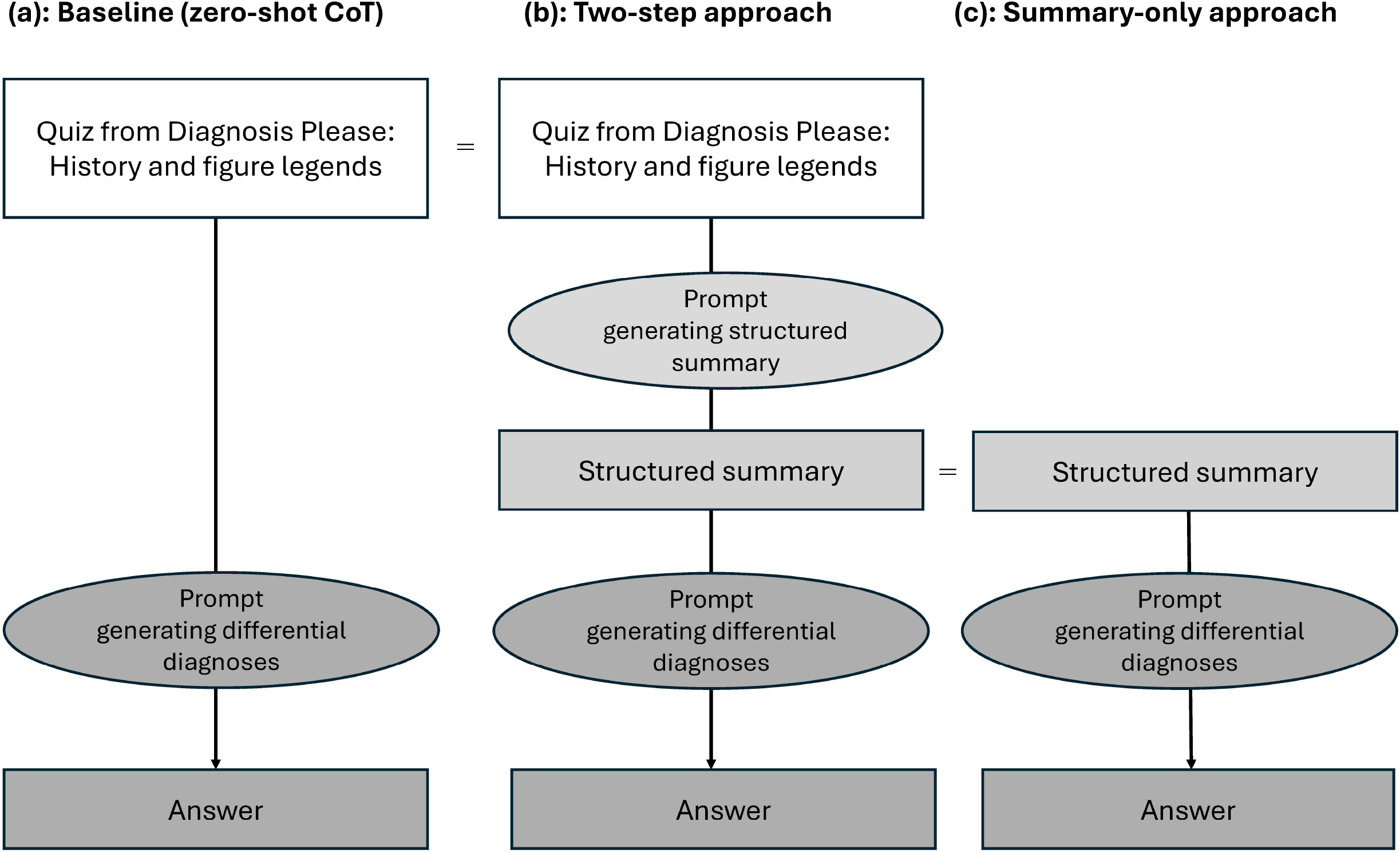
Proposed LLM workflow. (a) Baseline (zero-shot chain-of thought). (b) Two-step approach. (c). Summary-only approach. CoT: Chain of thought.

We compared three prompting approaches in this study. The baseline approach utilized a zero-shot chain-of-thought prompt with a role-play prompt (15), as was widely used in previous studies (9,12,13,16), designed to elicit a direct diagnostic response: “As a physician, I plan to utilize you for research purposes. Assuming you are a hypothetical physician, please walk me through the process from differential diagnosis to the most likely diagnosis and the next two most likely differential diagnoses step by step, based on the attached information.”

As the second approach, we utilized a two-step prompting strategy. The first step focused on information summarization, with the prompt: “You are an experienced Diagnostic Radiologist. Your task is to summarize the following clinical case, aiming to understand it thoroughly and determine the correct diagnosis. Categorize and summarize the information from the following clinical case into the specified categories. Use concise bullet points for each category, ensuring all critical information is captured. If a category has no relevant information, write ‘No information provided.” Categories: patient information (e.g., age, sex, race), history of present illness, past medical history, family history, social history (including relevant lifestyle factors), current medications and allergies, symptoms, physical examination findings, vital signs, laboratory results (highlight abnormalities), imaging findings, and additional relevant information”. The second step focused on diagnostic reasoning, using the prompt: “As a physician, I plan to utilize you for research purposes. Assuming you are a hypothetical physician, please walk me through the process from differential diagnosis to the most likely diagnosis and the next two most likely differential diagnoses step by step, based on the summarized information.”

As the third approach, without inputting the original text of the quiz case, we used only the summarized information obtained from the first step output in the second approach mentioned above as input, and had it infer the diagnosis in a separate session. We used the same prompt as the previous two-step prompting strategy.

One trainee radiologist and one board-certified diagnostic radiologist with 11 years of experience judged the correctness of LLM-generated most likely and differential diagnoses.

McNemar tests were used to evaluate the differences in accuracy rates for the top three differential diagnoses between the three methods. Two-sided p values < 0.05 were considered statistically significant. Statistical analyses were performed using the R software (version 4.1.1; R Foundation for Statistical Computing, Vienna, Austria). As this study utilized only published articles as data source, institutional review board approval was not required.

## Results

### The results are summarized in Table 1

The two-step approach demonstrated superior performance compared to both the baseline and summary-only methods. Examples of the LLM’s output are shown in Fig. 2. For primary diagnosis, the accuracy improved from 56.5% with the baseline approach to 60.6% with the two-step approach. This improvement was statistically significant (p = 0.042, McNemar test). The accuracy of the summary-only approach was 56.2%, which was significantly lower than the two-step approach (p = 0.035).

**Table 1:**
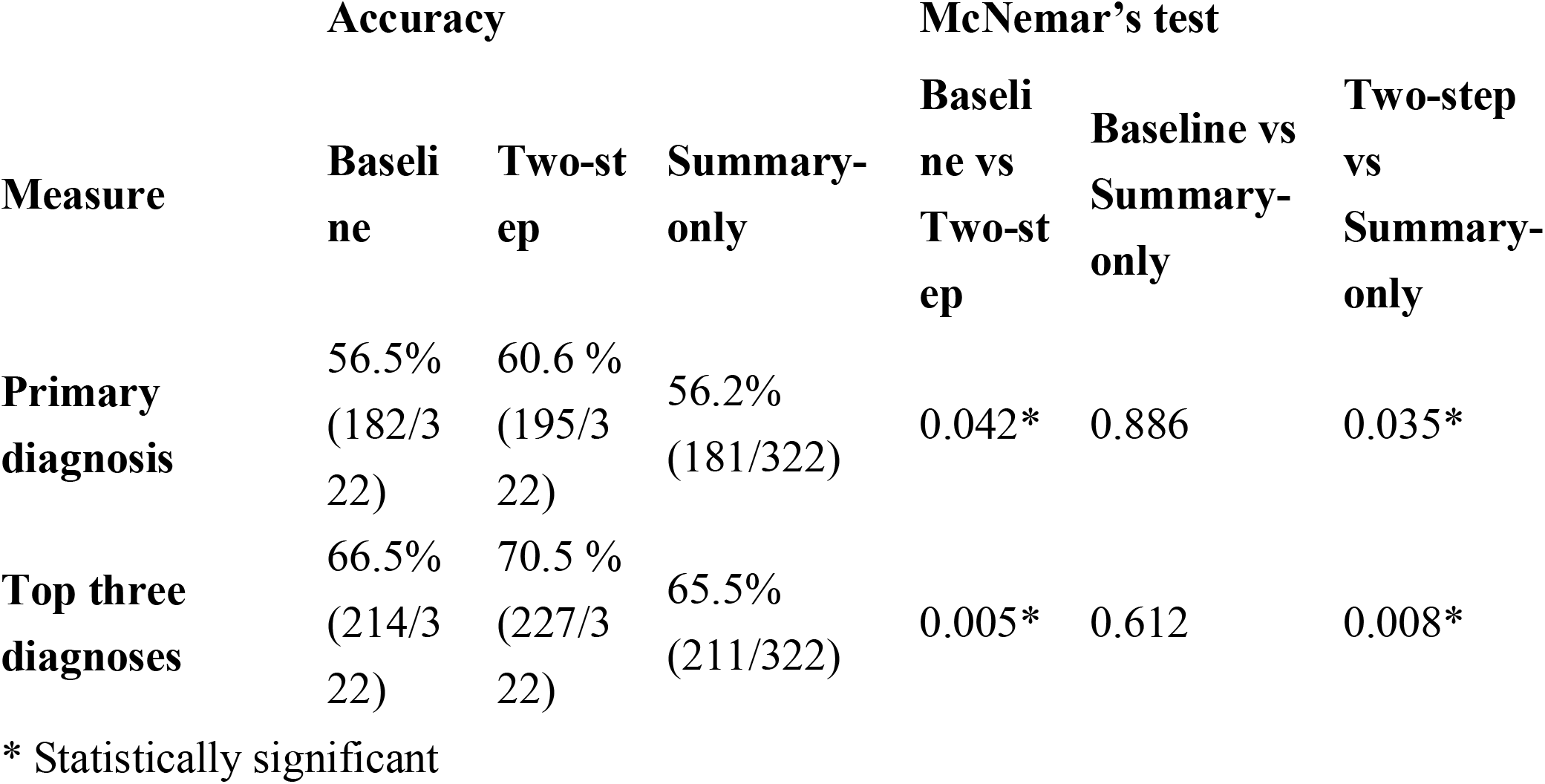
Comparison of Diagnostic Accuracy Across Methods and Results of McNemar’s Tests.

| Measure | Accuracy |  |  | McNemar's test |  |  |
| --- | --- | --- | --- | --- | --- | --- |
|  | Baseline | Two-step | Summary-only | Baseline vs Two-step | Baseline vs Summary-only | Two-step vs Summary-only |
| **Primary diagnosis** | 56.5%<br>(182/322) | 60.6 %<br>(195/322) | 56.2%<br>(181/322) | 0.042* | 0.886 | 0.035* |
| **Top three diagnoses** | 66.5%<br>(214/322) | 70.5 %<br>(227/322) | 65.5%<br>(211/322) | 0.005* | 0.612 | 0.008* |

\* Statistically significant

**Fig 2.**
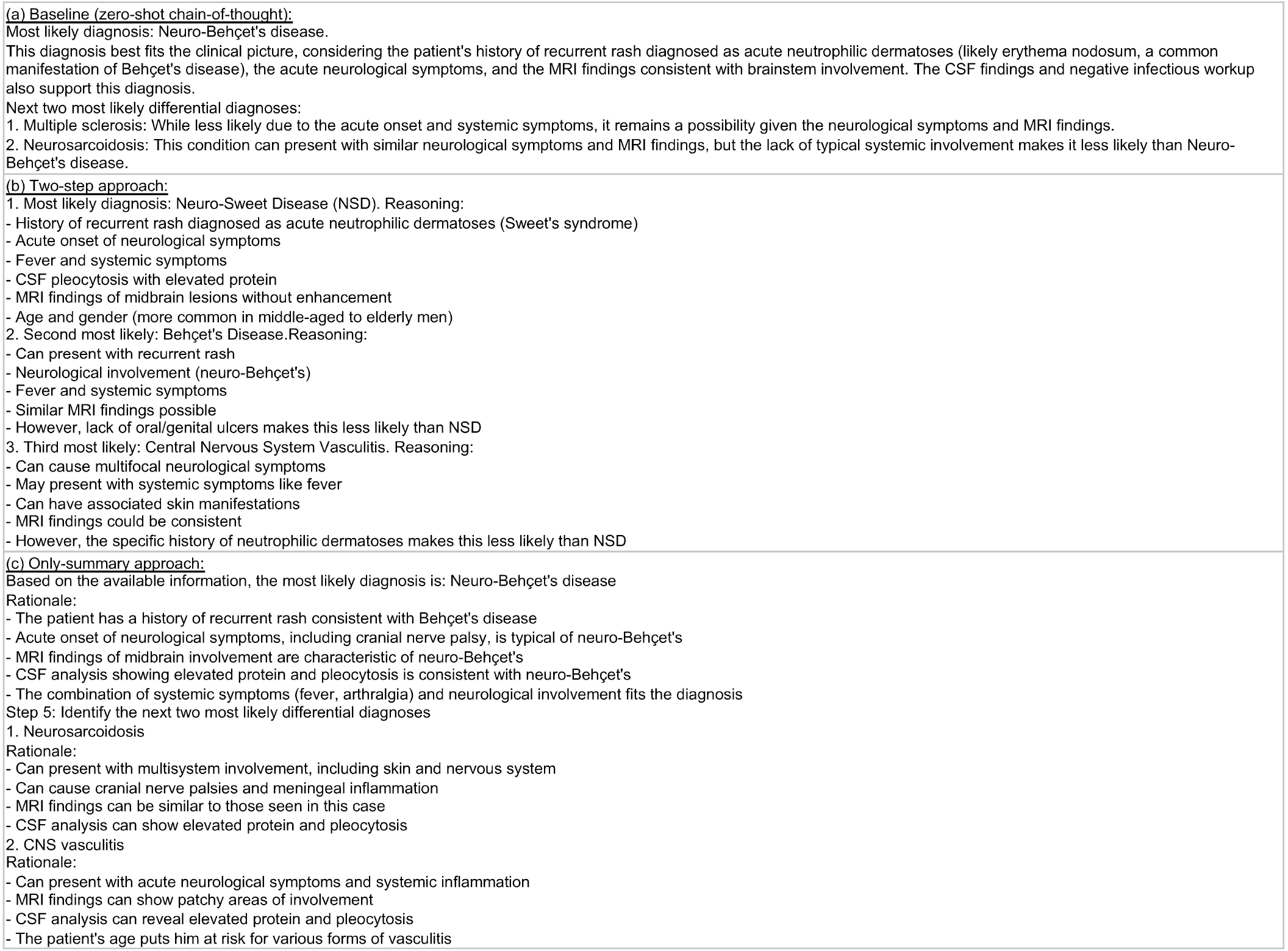
An example of the Claude 3 Sonnet-generated text for Case 176 whose correct diagnosis was “Neuro-Sweet Syndrome” (17). (a) A generated list of differential diagnoses for Case 176 using the conventional zero-shot chain-of-thought (baseline). (b) A generated list of differential diagnoses in the second step in the two-step approach. (c) A generated list of differential diagnoses in the summary-only approach. This case is an example where the two-step approach showed different conclusions than the other two approaches. In the two-step approach, the LLM made the correct diagnosis, neuro-Sweet’s disease, based on the patient’s symptoms, previous skin biopsy results of acute neutrophilic dermatosis, and imaging findings. It listed neuro-Behçet’s disease, which could present with similar progression and images, as the second differential diagnosis, because of lack of oral/genital ulcers. On the other hand, the other two approaches did not mention this and listed neuro-Behçet’s disease as the primary diagnosis, while failing to include neuro-Sweet’s disease in the differential diagnoses. The prompts used for Case 176, the generated summary, and the LLM’s outputs are provided in Supplementary Table 1.

For the top three diagnoses, the overall accuracy improved from 66.5% in the baseline to 70.5% with the two-step approach, demonstrating a statistically significant difference (p = 0.005). The summary-only approach achieved 65.5% accuracy for the top three diagnoses, which was also significantly lower than the two-step approach (p = 0.008). No significant differences were observed between the baseline and summary-only approaches for either primary diagnosis or top three diagnoses accuracy.

## Discussion

In this study, we compared the diagnostic ability of LLMs based on *Radiology’s* Diagnosis Please cases by having the LLM summarize the medical history in advance. We also examined the accuracy when only the summary obtained by this method was used as input, and compared it with the accuracy of the baseline and the two-step approach.

Our results demonstrated that incorporating LLM-generated structured clinical summaries at the midpoint significantly improved diagnostic accuracy compared to the baseline. The improved performance observed with our two-step approach could be attributed to several factors. First, the structured summarization step in our approach mirrors the process human clinicians use to organize and prioritize clinical information, incorporating traditional clinical knowledge summarization schemes that continue to be widely used in clinical reasoning today. Moreover, our findings align with research in other fields, suggesting that decomposing complex problems into simpler, manageable tasks often yields superior results (10). Our method of breaking down complex clinical problems into manageable, atomic problems or information might facilitate easier problem-solving, allowing for successful complex clinical reasoning and potentially suppressing errors.

A vital advantage of this two-step method is its applicability to virtually any situation requiring clinical reasoning only with minimal increase in computational complexity. The results of our two-step approach suggest the LLM’s potential to summarize and distill relevant information from free-text clinical data. In this study, even when only the summarized information was given to the LLM, although there was a slight decrease in accuracy compared to the baseline, no significant difference was observed. This aligns with previous research demonstrating LLMs’ proficiency in extracting and managing important clinical information from electronic health records (18,19,20). However, in the present results, the summary of the clinical history couldn’t fully replace the original free-text information in diagnosing patients’ disease. This suggests that there may have been information that the LLM couldn’t fully capture in its summary, or that presenting the patient’s history from multiple perspectives (i.e., both the original text and the summary) contributed to the LLM’s improved performance.

The structure of the summary used in this study was based on common clinical practice. However, this setup is arbitrary and may not be optimal for all cases. Future work could explore more flexible or case-specific structuring of clinical information. Additionally, our current approach leaves the summarization of imaging findings largely unstructured. Implementing a more structured approach to describing imaging results could potentially further improve diagnostic accuracy, especially in the field of diagnostic radiology.

Studies measuring LLM performance using publicly available journal quizzes always carry the potential for data leakage bias, where the input data itself may have been used in the LLM’s training. However, in this study, we input the same data into the baseline and the two-step methods, and thus the ultimate improvement observed in the two-step method can be considered a result of the method itself. Moreover, in the only-summary method, the structure of the input was substantially different from the original data, yet it achieved results nearly identical to the baseline. This suggests that the LLM is capable of clinical reasoning in an essential sense.

Several limitations of our study should be noted. First, the effectiveness of our method relies heavily on the LLM’s ability to accurately summarize clinical information. While Claude 3.5 Sonnet demonstrated strong capabilities in this regard, it may not generalize to all LLMs. Furthermore, the results may vary depending on the specific architecture, training dataset, and other inherent characteristics of the LLM, as well as the nature of the clinical reasoning problems used for comparison and the format in which these problems are presented. These factors could potentially lead to different outcomes across various scenarios and model implementations. Second, this study solely investigated solving medical quiz cases presented in text form. Therefore, future studies should test and refine this method using a variety of LLMs, datasets, and real-world scenarios.

## Conclusion

This study demonstrates the efficacy of a novel two-step approach in LLMs performance in medical diagnostics. Our findings suggest that this method, which aligns well with established clinical reasoning processes, could be valuable in utilizing LLMs in real-world clinical scenarios. The approach offers benefits with minimal drawbacks, potentially making it a useful tool for the practical implementation of LLMs in clinical decision support systems. Future research should focus on refining these techniques and exploring their applicability across diverse medical settings.

## Data Availability

All data produced in the present study are available upon reasonable request to the authors

