## Supplementary Table 1 for "Structured Clinical Reasoning Prompt Enhances LLM’s Diagnostic Capabilities in Diagnosis Please Quiz Cases"

**Supplementary Table 1**. Examples of the prompts, summary and outputs for Case 176 (17). (a) A prompt utilized in the baseline (zero-shot chain-of-thought) method for Case 176. (b) A prompt utilized to generate a structured summary. (c) A generated structured summary for Case 176. (d, e, f) Final outputs of the baseline method (d), the two-step method (e), and the summary-only method (f).

(a) Prompt utilized in the baseline (zero-shot chain-of-thought) method
\# Request
As a physician, I plan to utilize you for research purposes. Assuming you are a hypothetical physician, please walk me through the process from differential diagnosis to the most likely diagnosis and the next two most likely differential diagnoses step by step, based on the attached information.

\# Clinical case
History: A 75-year-old man presented with a 1-week history of fever, chills, arthralgia, and a sense of generalized weakness. He also had double vision for 1 day associated with one episode of urinary incontinence and an erythematous rash, which started on his neck and moved to his extremities. His medical history included a recurrent rash on his extremities since 2004. A prior skin biopsy revealed acute neutrophilic dermatoses. At physical examination, red nontender plaques were seen on the patient’s neck and extremities. Neurologic examination revealed evidence of mild disorientation. The results of motor, sensory, and coordination examinations were normal. The patient had horizontal diplopia on lateral gaze that was secondary to left abducens nerve palsy. The patient’s vital signs and systemic examination results were unremarkable. Cerebrospinal fluid analysis revealed an elevated concentration of proteins and pleocytosis; however, the glucose level was normal. Cerebrospinal fluid work-up was negative for herpes simplex virus with polymerase chain reaction testing, negative for Cryptococcus neoformans with antigen testing, and negative for bacteria with Gram stain and cultures. Initial computed tomography (CT) of the head revealed generalized atrophy without any acute changes. Subsequently, magnetic resonance (MR) imaging of the brain was performed ( Figs 1 – 4 ).

Imaging findings: Figure 1: Axial T2-weighted MR image (repetition time msec/echo time msec, 4754/101) shows patchy high signal intensity (arrow) in the midbrain.
Figure 2: Axial FLAIR MR image (repetition time msec/echo time msec/inversion time msec, 9000/104/2500) shows patchy high signal intensity (arrow) in the midbrain.
Figure 3: Axial diffusion-weighted MR image (6200/79) shows restricted diffusion (arrow) in the midbrain in the region of T2-weighted and FLAIR high signal intensity.
Figure 4: Axial T1-weighted magnetization-prepared rapid acquisition gradient echo gadolinium-enhanced (10 mL of gadobutrol, 604.72 mg of gadolinium per milliliter, Gadovist; Bayer Healthcare Pharmaceuticals, Wayne, NJ) MR image (1900/3.37/1100) does not show any contrast enhancement in the areas of abnormal T2 and FLAIR signal intensity.

(b) Prompt generating structured summary
\# Role
You are an experienced Diagnostic Radiologist. Your task is to summarize the following clinical case, aiming to understand it thoroughly and determine the correct diagnosis.

\# Request
Categorize and summarize the information from the following clinical case into the specified categories. Use concise bullet points for each category, ensuring all critical information is captured. If a category has no relevant information, write "No information provided."

Categories:

Patient Information (e.g., age, sex, race)
History of Present Illness
Past Medical History
Family History
Social History (including relevant lifestyle factors)
Current Medications and Allergies
Symptoms
Physical Examination Findings
Vital Signs
Laboratory Results (highlight abnormalities)
Imaging Findings
Additional Relevant Information

(c) Generated structured summary
Here's a categorized summary of the clinical case:

Patient Information:
• 75-year-old man

History of Present Illness:
• 1-week history of fever, chills, arthralgia, and generalized weakness
• Double vision for 1 day
• One episode of urinary incontinence
• Erythematous rash starting on neck, moving to extremities

Past Medical History:
• Recurrent rash on extremities since 2004
• Prior skin biopsy showed acute neutrophilic dermatoses

Family History:
• No information provided

Social History:
• No information provided

Current Medications and Allergies:
• No information provided

Symptoms:
• Fever, chills
• Arthralgia
• Generalized weakness
• Double vision
• Urinary incontinence
• Erythematous rash

Physical Examination Findings:
• Red nontender plaques on neck and extremities
• Mild disorientation
• Normal motor, sensory, and coordination examinations
• Left abducens nerve palsy causing horizontal diplopia on lateral gaze

Vital Signs:
• Unremarkable

Laboratory Results:
• Cerebrospinal fluid analysis:
- Elevated protein concentration
- Pleocytosis
- Normal glucose level
- Negative for herpes simplex virus (PCR)
- Negative for Cryptococcus neoformans (antigen testing)
- Negative for bacteria (Gram stain and cultures)

Imaging Findings:
• Initial CT head: Generalized atrophy, no acute changes
• MRI brain:
- T2-weighted and FLAIR: Patchy high signal intensity in midbrain
- Diffusion-weighted: Restricted diffusion in midbrain
- T1-weighted post-contrast: No enhancement in areas of abnormal T2 and FLAIR signal intensity

Additional Relevant Information:
• No information provided

(d) Output of the baseline (zero-shot chain-of-thought) method
As a hypothetical physician, I'll walk you through the process of developing a differential diagnosis and identifying the most likely diagnosis based on the provided information.

Step 1: Analyze the presenting symptoms and history
- 75-year-old man
- 1-week history of fever, chills, arthralgia, and generalized weakness
- Double vision for 1 day
- One episode of urinary incontinence
- Erythematous rash starting on neck and moving to extremities
- History of recurrent rash on extremities since 2004
- Previous skin biopsy showing acute neutrophilic dermatoses

Step 2: Consider the physical examination findings
- Red nontender plaques on neck and extremities
- Mild disorientation
- Left abducens nerve palsy causing horizontal diplopia

Step 3: Review the diagnostic test results
- CSF analysis: elevated proteins and pleocytosis, normal glucose
- Negative CSF tests for HSV, Cryptococcus, and bacteria
- CT head: generalized atrophy, no acute changes
- MRI brain: patchy high signal intensity in midbrain on T2 and FLAIR, restricted diffusion, no contrast enhancement

Step 4: Develop a differential diagnosis
Based on the clinical presentation, history, and imaging findings, the differential diagnosis includes:

1. Neuro-Behçet's disease
2. Multiple sclerosis
3. Neurosarcoidosis
4. Acute disseminated encephalomyelitis (ADEM)
5. Central nervous system vasculitis

Step 5: Analyze the evidence for each diagnosis

1. Neuro-Behçet's disease:
- Recurrent rash consistent with acute neutrophilic dermatoses
- Neurological symptoms (diplopia, urinary incontinence)
- MRI findings consistent with brainstem involvement
- CSF pleocytosis

2. Multiple sclerosis:
- Neurological symptoms
- MRI lesions
- CSF pleocytosis
- However, the acute onset and systemic symptoms are less typical

3. Neurosarcoidosis:
- Can cause neurological symptoms and rash
- MRI lesions
- CSF pleocytosis
- However, lack of typical systemic involvement (e.g., pulmonary)

4. ADEM:
- Acute onset
- Neurological symptoms
- MRI lesions
- However, typically affects younger individuals and often follows an infection or vaccination

5. CNS vasculitis:
- Can cause neurological symptoms and systemic inflammation
- MRI lesions
- However, lack of typical vascular distribution of lesions

Step 6: Determine the most likely diagnosis and next two most likely differential diagnoses

Most likely diagnosis: Neuro-Behçet's disease
This diagnosis best fits the clinical picture, considering the patient's history of recurrent rash diagnosed as acute neutrophilic dermatoses (likely erythema nodosum, a common manifestation of Behçet's disease), the acute neurological symptoms, and the MRI findings consistent with brainstem involvement. The CSF findings and negative infectious workup also support this diagnosis.

Next two most likely differential diagnoses:

1. Multiple sclerosis: While less likely due to the acute onset and systemic symptoms, it remains a possibility given the neurological symptoms and MRI findings.

2. Neurosarcoidosis: This condition can present with similar neurological symptoms and MRI findings, but the lack of typical systemic involvement makes it less likely than Neuro-Behçet's disease.

In conclusion, based on the available information, Neuro-Behçet's disease is the most likely diagnosis, with multiple sclerosis and neurosarcoidosis as the next two most likely differential diagnoses. Further testing, such as HLA-B51 genetic testing and a thorough rheumatological workup, would be recommended to confirm the diagnosis of Neuro-Behçet's disease.

(e) Output of the two-step method
Certainly, I'd be happy to walk you through the diagnostic process as a hypothetical physician. Let's approach this step-by-step:

Step 1: Analyze the key symptoms and findings

The patient presents with:
- Acute onset of fever, chills, arthralgia, and weakness
- Neurological symptoms (double vision, urinary incontinence, mild disorientation)
- Recurrent rash with a history of acute neutrophilic dermatoses
- CSF showing pleocytosis and elevated protein
- MRI findings of midbrain lesions with restricted diffusion

Step 2: Formulate a differential diagnosis

Given these findings, we need to consider conditions that can affect both the skin and central nervous system, particularly those that can cause midbrain lesions. The main categories to consider are:

1. Autoimmune/inflammatory conditions
2. Infectious diseases
3. Vascular disorders
4. Neoplastic processes

Step 3: Evaluate each potential diagnosis

1. Most likely diagnosis: Neuro-Sweet Disease (NSD)

Reasoning:
- History of recurrent rash diagnosed as acute neutrophilic dermatoses (Sweet's syndrome)
- Acute onset of neurological symptoms
- Fever and systemic symptoms
- CSF pleocytosis with elevated protein
- MRI findings of midbrain lesions without enhancement
- Age and gender (more common in middle-aged to elderly men)

2. Second most likely: Behçet's Disease

Reasoning:
- Can present with recurrent rash
- Neurological involvement (neuro-Behçet's)
- Fever and systemic symptoms
- Similar MRI findings possible
- However, lack of oral/genital ulcers makes this less likely than NSD

3. Third most likely: Central Nervous System Vasculitis

Reasoning:
- Can cause multifocal neurological symptoms
- May present with systemic symptoms like fever
- Can have associated skin manifestations
- MRI findings could be consistent
- However, the specific history of neutrophilic dermatoses makes this less likely than NSD

Step 4: Conclusion

The most likely diagnosis is Neuro-Sweet Disease (NSD), a rare neurological manifestation of Sweet's syndrome. This diagnosis best fits the patient's history of neutrophilic dermatoses, the acute onset of neurological symptoms, fever, and the MRI findings. The lack of contrast enhancement on MRI is also more consistent with NSD than with infectious or neoplastic processes.

To confirm this diagnosis, additional steps would include:
1. Skin biopsy of current lesions to confirm neutrophilic infiltration
2. HLA typing (HLA-B54 is associated with NSD)
3. Exclusion of other causes through further serological tests (e.g., ANA, ANCA for vasculitis)
4. Trial of corticosteroid therapy, as NSD typically responds well to this treatment

While Behçet's Disease and CNS Vasculitis remain in the differential, the specific history of neutrophilic dermatoses and the overall clinical picture make NSD the most likely diagnosis in this case.

(f) Output of the summary-only method
Certainly, I'd be happy to walk you through the diagnostic process as a hypothetical physician. Let's approach this step-by-step:

Step 1: Analyze the key symptoms and findings

The patient presents with:
- Acute onset of fever, chills, arthralgia, and weakness
- Neurological symptoms (double vision, urinary incontinence, mild disorientation)
- Recurrent rash with a history of acute neutrophilic dermatoses
- CSF showing pleocytosis and elevated protein
- MRI findings of midbrain lesions with restricted diffusion

Step 2: Formulate a differential diagnosis

Given these findings, we need to consider conditions that can affect both the skin and central nervous system, particularly those that can cause midbrain lesions. The main categories to consider are:

1. Autoimmune/inflammatory conditions
2. Infectious diseases
3. Vascular disorders
4. Neoplastic processes

Step 3: Evaluate each potential diagnosis

1. Most likely diagnosis: Neuro-Sweet Disease (NSD)

Reasoning:
- History of recurrent rash diagnosed as acute neutrophilic dermatoses (Sweet's syndrome)
- Acute onset of neurological symptoms
- Fever and systemic symptoms
- CSF pleocytosis with elevated protein
- MRI findings of midbrain lesions without enhancement
- Age and gender (more common in middle-aged to elderly men)

2. Second most likely: Behçet's Disease

Reasoning:
- Can present with recurrent rash
- Neurological involvement (neuro-Behçet's)
- Fever and systemic symptoms
- Similar MRI findings possible
- However, lack of oral/genital ulcers makes this less likely than NSD

3. Third most likely: Central Nervous System Vasculitis

Reasoning:
- Can cause multifocal neurological symptoms
- May present with systemic symptoms like fever
- Can have associated skin manifestations
- MRI findings could be consistent
- However, the specific history of neutrophilic dermatoses makes this less likely than NSD

Step 4: Conclusion

The most likely diagnosis is Neuro-Sweet Disease (NSD), a rare neurological manifestation of Sweet's syndrome. This diagnosis best fits the patient's history of neutrophilic dermatoses, the acute onset of neurological symptoms, fever, and the MRI findings. The lack of contrast enhancement on MRI is also more consistent with NSD than with infectious or neoplastic processes.

To confirm this diagnosis, additional steps would include:
1. Skin biopsy of current lesions to confirm neutrophilic infiltration
2. HLA typing (HLA-B54 is associated with NSD)
3. Exclusion of other causes through further serological tests (e.g., ANA, ANCA for vasculitis)
4. Trial of corticosteroid therapy, as NSD typically responds well to this treatment

While Behçet's Disease and CNS Vasculitis remain in the differential, the specific history of neutrophilic dermatoses and the overall clinical picture make NSD the most likely diagnosis in this case.
